# GestaltGAN: Synthetic photorealistic portraits of individuals with rare genetic disorders

**DOI:** 10.1101/2024.07.18.24308205

**Authors:** Aron Kirchhoff, Alexander Hustinx, Behnam Javanmardi, Tzung-Chien Hsieh, Fabian Brand, Shahida Moosa, Thomas Schultz, Benjamin D. Solomon, Peter Krawitz

## Abstract

The facial gestalt (overall facial morphology) is a characteristic clinical feature in many genetic disorders that is often essential for suspecting and establishing a specific diagnosis. For that reason, publishing images of individuals affected by pathogenic variants in disease-associated genes has been an important part of scientific communication. Furthermore, medical imaging data is also crucial for teaching and training artificial intelligence methods such as GestaltMatcher. However, medical data is often sparsely available and sharing patient images involves risks related to privacy and re-identification. Therefore, we explored whether generative neural networks can be used to synthesize accurate portraits for rare disorders. We modified a StyleGAN architecture and trained it to produce random condition-specific portraits for multiple disorders. We present a technique that generates a sharp and detailed average patient portrait for a given disorder. We trained our GestaltGAN on the 20 most frequent disorders from the GestaltMatcher database. We used REAL-ESRGAN to increase the resolution of portraits from the training data with low quality and colorized black-and-white images. The training data was aligned and cropped to achieve a uniform format. To augment the model’s understanding of human facial features, an unaffected class was introduced to the training data.

We tested the validity of our generated portraits with 63 human experts. Our findings demonstrate the model’s proficiency in generating photorealistic portraits that capture the characteristic features of a disorder but preserve the patient’s privacy. Overall, the output from our approach holds promise for various applications, including visualizations for publications, educational materials, as well as augmenting training data for deep learning.

## Introduction

Many genetic conditions involve features that are evident on physical examination, including those that affect the face. At the time of writing (May 28, 2024), searching with the HPO term “facial dysmorphism” yields 2,997 entries in the Online Mendelian Inheritance of Men (OMIM) compendium (https://www.omim.org/), indicating the importance of the facial gestalt for characterizing disease entities. The importance of phenotype matching extends to diagnostic procedures in genetics, where physical examination features can serve as supporting evidence when assessing sequence variants for pathogenicity (Richards et al., 2015).

Recent advancements in computer vision have achieved expert-level accuracy in discerning distinct facial patterns. Next-generation phenotyping (NGP) tools such as GestaltMatcher have become instrumental in the analysis of clinical patterns in human faces and their usage for the interpretation of sequencing data (Hsieh, Bar-Haim et al., 2022; Hsieh, Mensah et al., 2019; Schmidt et al., 2023). The underlying technology, which is a deep convolutional neural network, can be used for pattern recognition as well as the delineation of informative features (explainable AI, XAI) or the synthesis of images with similar characteristics via generative methods such as Generative Neural Networks (GNN) (Saranya et al. 2023). GNNs may be particularly useful in medical settings since data are often sparsely available and may involve sensitive, private information. The generated images can be used for teaching or data augmentation when training machine learning models, including to address privacy concerns (Bowles et al. 2018, Shorten et al. 2019). In medical genetics, Duong, *et al*. showed that a StyleGAN can be used to generate artificial longitudinal data of patients, and could improve NGP classification accuracy by a better control of age as a confounder.^1^

StyleGAN is now a well-established architecture for image generation (Karras, Laine, et al., 2019), that allows the synthesis of photorealistic images across diverse contexts. StyleGAN is based on the concept of Generative Adversarial Networks originally proposed by Goodfellow et al. (2014), which consists of two parts, the generator and the discriminator. The generator crafts images—such as human portraits—while the discriminator evaluates their quality, providing feedback to reduce artifacts and enhance realism. The generator’s goal is to produce images so realistic that the discriminator cannot tell whether they are real or synthetic. Through this adversarial process, the generator learns characteristic object properties that are required to produce realistic synthetic images (in our case, human faces). A more comprehensive introduction to the technology can be found in the supplemental material (Related work).

With further refinement of GANs, it is also possible to condition the output depending on an input label (Mirza et al., 2014). This label is an additional piece of information that enables one to conditionally generate a certain type of image, in case of human faces the label might encode age, race or ethnicity, hair color, or even a certain genetic condition, as is the focus of our work.

Artificial content creation is particularly compelling in medicine given the sparse availability and stringent privacy constraints on data. However, facial images are also a particularly sensitive type of medical data, as the effort required for re-identification is relatively low and may require no additional technology. Nevertheless, for this study, the characteristics that are most suitable for de-identification can only be those that are not disease-related. There are therefore limits to anonymization in so far as recognition of the disease is our aim (k-anonymity is bound by the prevalence of the disorder). However, sparse training data poses challenges, potentially leading to overfitting, a phenomenon where the network memorizes samples and recreates training images (Shorten et al, 2019). Balancing de-identification with feature retention poses a nuanced challenge; the model must learn and reproduce disorder-specific features without replicating exact facial combinations from training images.

In our work, we used “disorder” as an additional class label and trained a conditional StyleGAN with images of the GestaltMatcher database (GMDB), which contains images of over 10,000 individuals with molecularly confirmed diagnoses (Lesmann et al. 2024).^2^ We hypothesized that working with several syndromic disorders facilitates learning certain clinical features that are often shared by more than one disorder.^2^ We therefore, focused on the 20 most frequent syndromes represented in the GMDB, comprising a total number of 3209 images. In order to enrich the characteristic features, we added a custom loss to the training and penalized the model if GestaltMatcher-Arc’s (Hustinx, et al., 2023) feedback would not match with the syndrome requested of our GAN.

For the evaluation of the generated images, we tested whether humans are still able to differentiate between synthetic and original images, and whether the characteristic features of a genetic condition are conserved, while data of real patients is protected.

## Methods and Materials

### Data Preparation

The GestaltMatcher DataBase (GMDB) contains a collection of 581 distinct disorders known to involve facial dysmorphisms, with over 10,980 accompanying images of 8346 affected individuals. In addition to the use of previously published images, all individuals newly represented in GMDB provided consent for using their imaging data for machine learning. For our GestaltGAN model training, we focused on the 20 most common disorders from the GMDB (Supplemental Figure 1). The reason for choosing 20 is a trade-off, balancing the benefit of using more disorders for training data against the challenge of distinguishing between them. Images in the GMDB come in various formats, with differences in size, lighting, and facial alignment. To enhance the quality of low-resolution images, we used REAL-ESRGAN (Wang et al., 2021), a deep learning model that predicts high-quality image details and computes high-resolution versions for the input images. Additionally, many older images in the GMDB come in black-and-white, which would lead to undesired outputs if directly used for training. To address this, we utilized DDColor (Kang, Yang et al., 2023) to add color to these monochromatic images, ensuring a consistent dataset for our GAN training. All images were aligned and cropped using GestaltEngine-FaceCropper, which relies on RetinaFace (Deng, Guo, Yuxiang, et al., 2019) that accurately pinpoints five landmark points in each portrait: the eyes, nose, and corners of the mouth. Using those landmarks, horizontal alignment of the faces could be ensured.

Given the scarcity of images of individuals with the genetic conditions of interest, we expanded our dataset by including images of individuals without known genetic conditions. These unaffected faces share similar features, like hair or skin, with the images of individuals with genetic conditions, aiding the model in generating more realistic faces of individuals with genetic conditions. We opted for the FFHQ-Aging dataset (Or-El et al., 2020), known for its size and high-quality images across different ages and races and ethnicities. Since many patients in the GMDB are children (with 42.8% under five years old), we balanced the age distribution by limiting the number of unaffected adults in FFHQ-Aging to 3000 individuals for all age groups over 20 years old. This adjustment resulted in a training set with 31,130 unaffected people. Subsequently, all images in FFHQ-Aging underwent alignment and cropping using GestaltEngine-FaceCropper.

### Training of GestaltGAN

We utilized the conditional StyleGAN3-R architecture as proposed by Karras, Aittala, Laine, et al. (2021), representing the fourth iteration of the StyleGAN framework. This version incorporates enhancements such as translation and rotation invariance towards training images, which helps with imperfect alignment that may have persisted in some training images. Notably, StyleGAN3-R integrates adaptive discriminator augmentation (ADA), a mechanism crucial for preventing overfitting, especially in datasets with limited samples like the GMDB.

Our approach is to use a conditional setup, where each syndrome is treated as a distinct class for training. This way, we can make the most of the shared facial features among different disorders, making use of the size of our own dataset. We also include unaffected images as a separate class, providing the model with continuous exposure to common features like hair or skin from those images. However, to handle the variable number of training images for each genetic condition, and to avoid producing images that inappropriately incorporate features of unaffected faces we added an over-sampling function to the StyleGAN3 implementation. This ensures that each genetic condition is represented equally during training. We gave the model twenty times more exposure to the unaffected class to allow the model to better understand and utilize its features.

In addition to over-sampling, we modified the loss function of StyleGAN3. Since the GestaltMatcher model is specialized at identifying rare disorders from images, we want to leverage this skill by penalizing our model if its predictions deviate from what GestaltMatcher would expect for a given class. The adjusted loss function combines this GestaltMatcher loss with the regular discriminator loss *L*_*D*_:

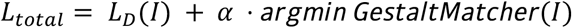

Here, *I* represents the image generated by the generator, and the GestaltMatcher function calculates the sequence of most likely diagnoses in ascending order. Argmin returns the index of the correct diagnosis. The weight *α* adjusts the balance between the GestaltLoss and the discriminator loss.

The chosen image resolution for our model was 256×256 pixels, which is slightly below the median image resolution in the GMDB of 265×328pixels. We were also able to train the model for a 512×512 resolution, but did not continue this approach due to the three-fold higher required computation effort. A visualization for our training setup is shown in Figure 2. Images generated by our GestaltGAN model can be seen in Figure 1.

**Fig. 1:**
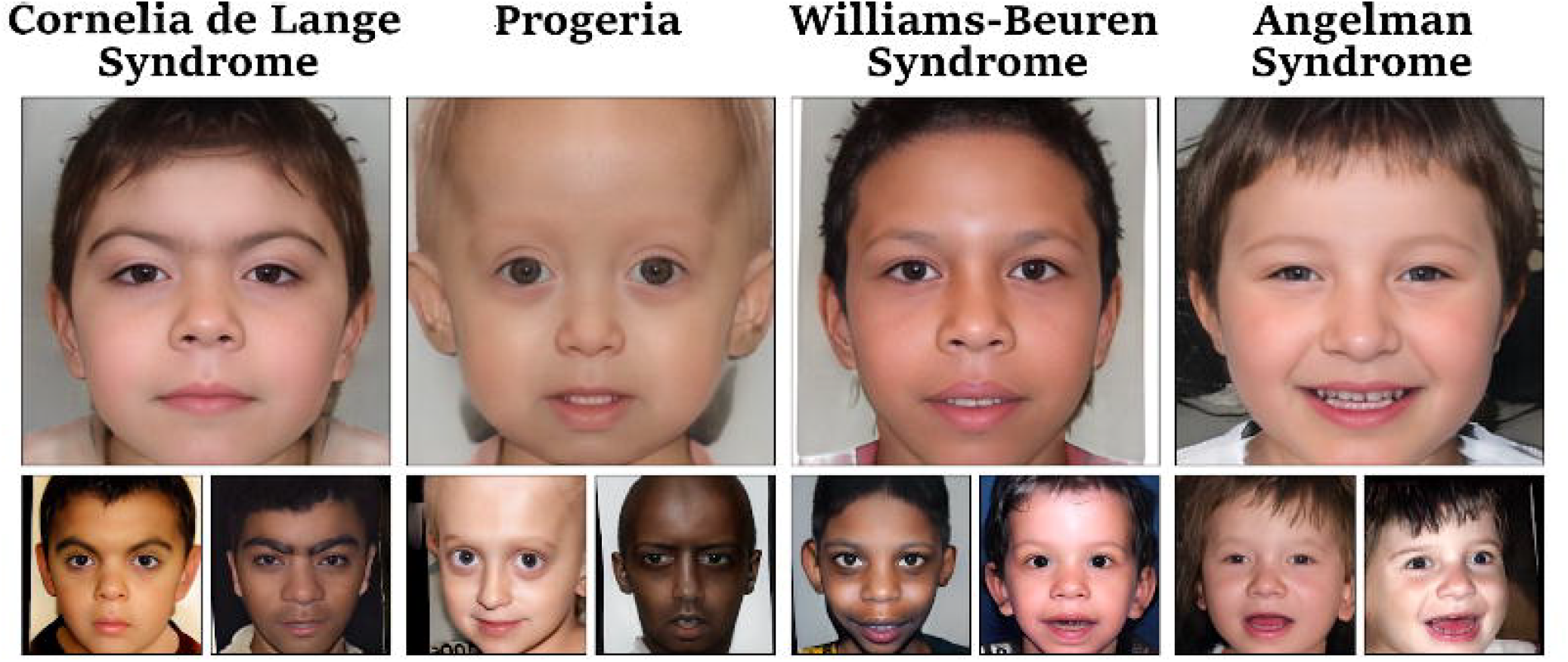
Images generated by GestaltGAN. Images in the top row are the latent representation for the disorder. in the bottom row are selected images generated for the respective disorder.

**Fig. 2:**
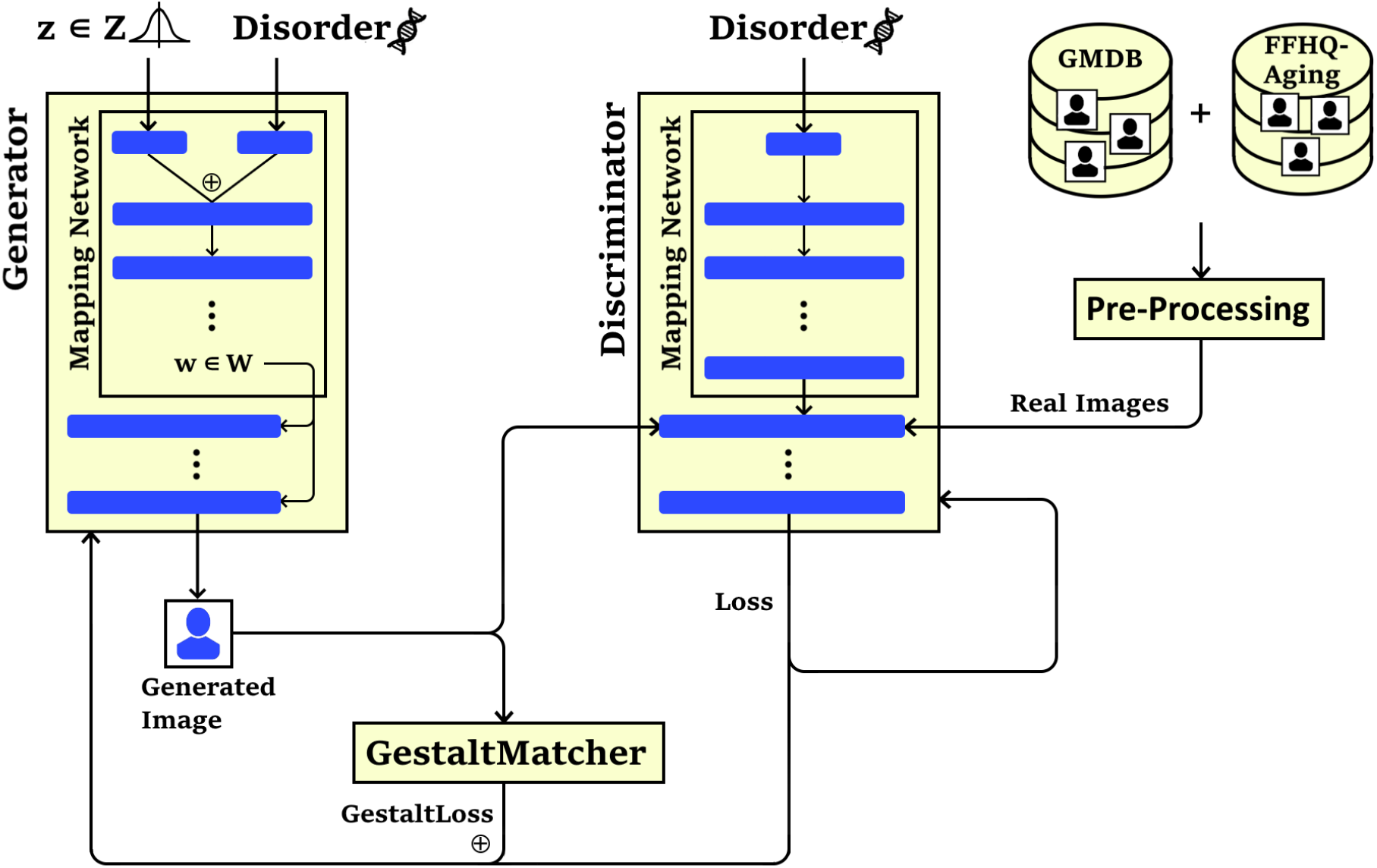
Visualization for the GestaltGAN architecture. StyleGAN has been extended by a customized loss function, GestaltLoss, based on the GestaltMatcher ensemble. The conditional generator synthesizes images for 20 different disorders and receives feedback from the discriminator about the origin, which is either artificial or real. For training of human faces and disorders, data of FFHQ Aging and the GMDB were used.

### Image averages and latent averages

To illustrate the characteristics of a disorder, the average face of several patients is often computed by registering and overlaying their portraits. While this method has been used in various studies, the resulting images are often blurry and indistinct^3^. In fact, increasing the number of individuals often leads to a deterioration of the results (personal communications). In this subsection, we introduce a technique to generate sharp and high-quality portraits that accurately represent the features of specific disorders.

The image generation process of an image with StyleGAN begins with sampling a random latent vector *z* ∈ *Z*. This vector is then combined with the class label in the mapping network, which maps to the second latent space *W*. An image is then deterministically generated based on this latent vector in *w* ∈ *W*. Since the latent space is continuous, small variations in the latent vector result in slight variations in the synthesized image (Xia et al., 2022). This means that similar images and especially images of the same disorder lie in the same region in the latent space. This property allows us to generate an average image for all disorders on which our model was trained. To achieve this, we sample 10,000 latent vectors *w* ∈ *W* for the selected disorder, such as Cornelia de Lange syndrome, and average these latent vectors to generate the average image. Instead of computing the average in the image space, we perform the averaging in the latent or feature space. Latent averages for different disorders are shown in Figure 1 and Figure 3. In Figure 3, the latent averages are presented alongside the corresponding averages from image space, which are currently often used for teaching purposes, such as to help clinical trainees recognize different genetic conditions.

**Fig. 3.**
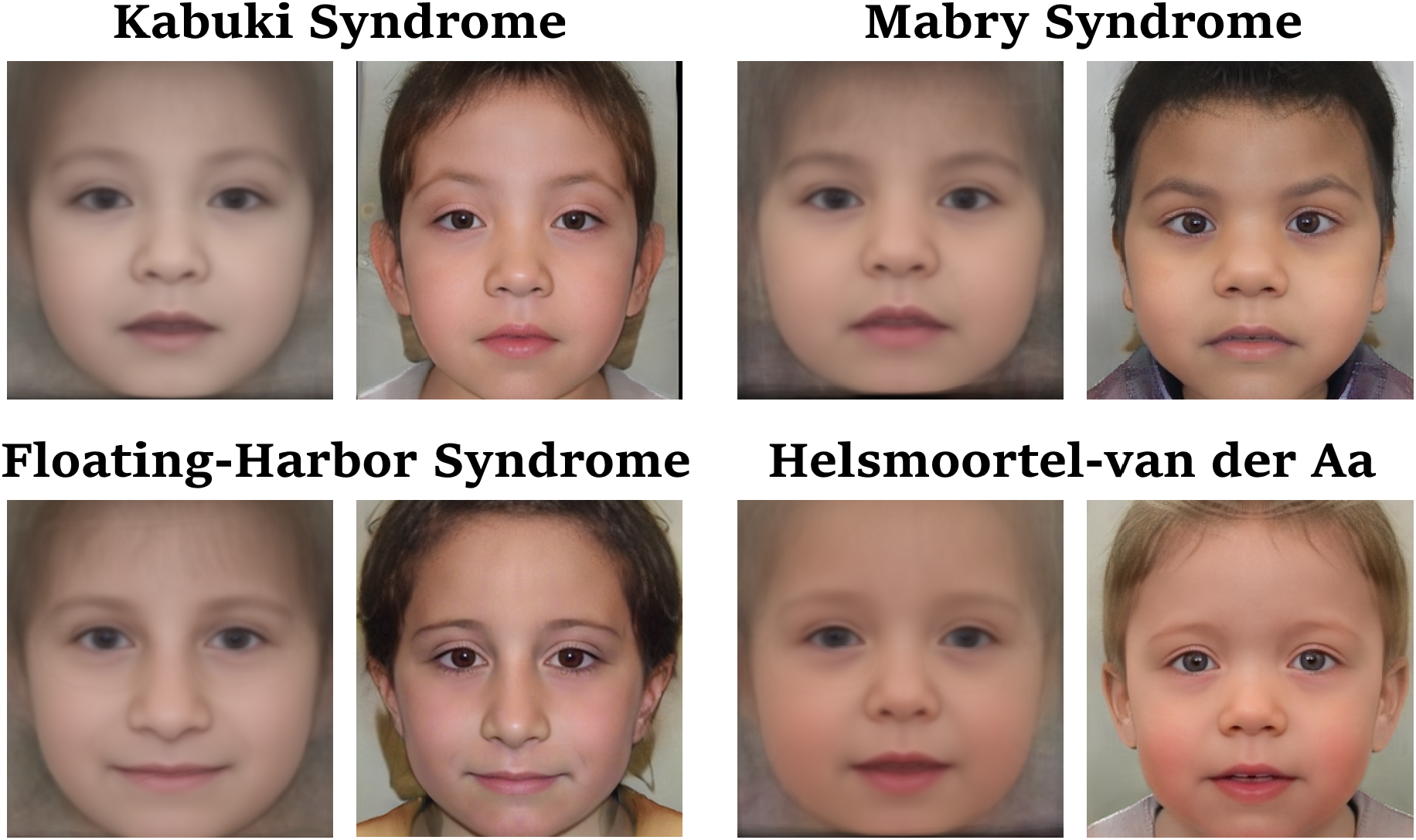
Comparison of ordinary image averages and latent averages. Since both averaging techniques operate in essence on the same underlying data, there is a high similarity of image averages (left) and latent averages (right) for each condition. However, averaging in image space blurs fine structures, while latent averages appear more photorealistic.

An important note is that this method only works for disorders on which the GAN has been. To extend the capability and generate latent averages for other disorders, we perform GAN-inversion on all patient portraits in the cohort to obtain the corresponding feature vectors in the latent space that represents the patients. We use the GestaltMatcher ensemble as a loss function for GAN inversion, as it has been trained to recognize dysmorphic features. After averaging the feature vectors, we generate an image from the resulting vector to obtain the latent average.

## Results

With GestaltGAN, our goals were threefold: we aimed to create synthetic images that are photorealistic, de-identified, and which accurately represent clinical features. Achieving these objectives partially involved navigating an optimization problem, as enhancing privacy protection might sometimes compromise feature preservation. We therefore assessed the quality of all three objectives through computational methods and by conducting experimental evaluations with human test participants, who compared the generated images to the original images. Images generated by our model are shown in Figure 1.

### Computational evaluation of image quality

To evaluate the quality of a large quantity of generated images, we employed two machine learning-based methods. First, we aimed to determine whether an image depicts a face or is considered a fail case, and second, we assessed whether characteristic features of the disorders are present in the images.

We generated 1000 random images for each class, including the unaffected faces class. While most generated images are high-quality portraits, some fail to convey meaningful content. We defined a fail-case as an image without any visible face. To estimate the number of fail-cases, we utilized RetinaFace, which was already used for image alignment. Since RetinaFace predicts facial features like eyes, nose, and mouth, its accuracy can be considered to represent a sensible criterion; we considered an image a fail-case if the confidence of RetinaFace was below 99.9% (Supplemental Figure 2). The percentage of fail-cases is below 10% for most disorders, but the proportion of fail-cases varies between disorders, such as 2.7% and 6.9% for Cornelia de Lange syndrome and Kabuki syndrome, respectively (Supplemental Figure 3). Possible reasons for this could include lower training image quality or more unique facial features in certain disorders. Unique features pose a challenge for the model, as they are encountered less frequently in the training data.

To assess whether characteristic phenotypes of the disorders are represented in the generated images, we utilized GestaltMatcher. We tested whether the generated disorder was within its top-5 predictions. Overall, GestaltMatcher achieved a top-5 accuracy of 76.7% on the synthesized images. The top-5-accuracy rates for Cornelia de Lange syndrome and Williams-Beuren syndrome were above 90%, while the correct disorder was only listed in the top 5 differential diagnoses for synthetic images of Baraitser-Winter in 62.1% and for Nicolaides-Baraitser in 39.9%. These performances are in good agreement with the accuracy rates measured on real data, in which top accuracy rates also differ per disorder depending on their distinctiveness. Therefore, the results indicate that the characteristic features are indeed present in the generated images.

### Assessment of image quality by human experts

In addition to the computational techniques, we used to assess image quality, we also developed an online survey with questions in three categories. We invited users of the GestaltMatcher databas to participate in the survey. These users represent dysmorphologists and other medical professionals working on genetic and other rare disorders, The experiment enabled us to assess the performance of humans in distinguishing 1) synthetic images from non-synthetic images, 2) re-identifying original data (images) that were used for training, and 3) identifying (diagnosing) the correct disorder. Each survey question also included a time-limit to prevent participants from scanning images for tiny artifacts, which can occur in synthetic images. In addition, there was a skip button that allowed participants to skip a question and continue with the next question. In total we recorded 63 sessions, in which 10 questions in each of the three categories were asked. Out of the 1860 answers we excluded 106 skipped questions and 151 timeouts resulting in 1603 answers for further evaluations. The full experiment setup is visualized in Figure 4.

**Figure 4:**
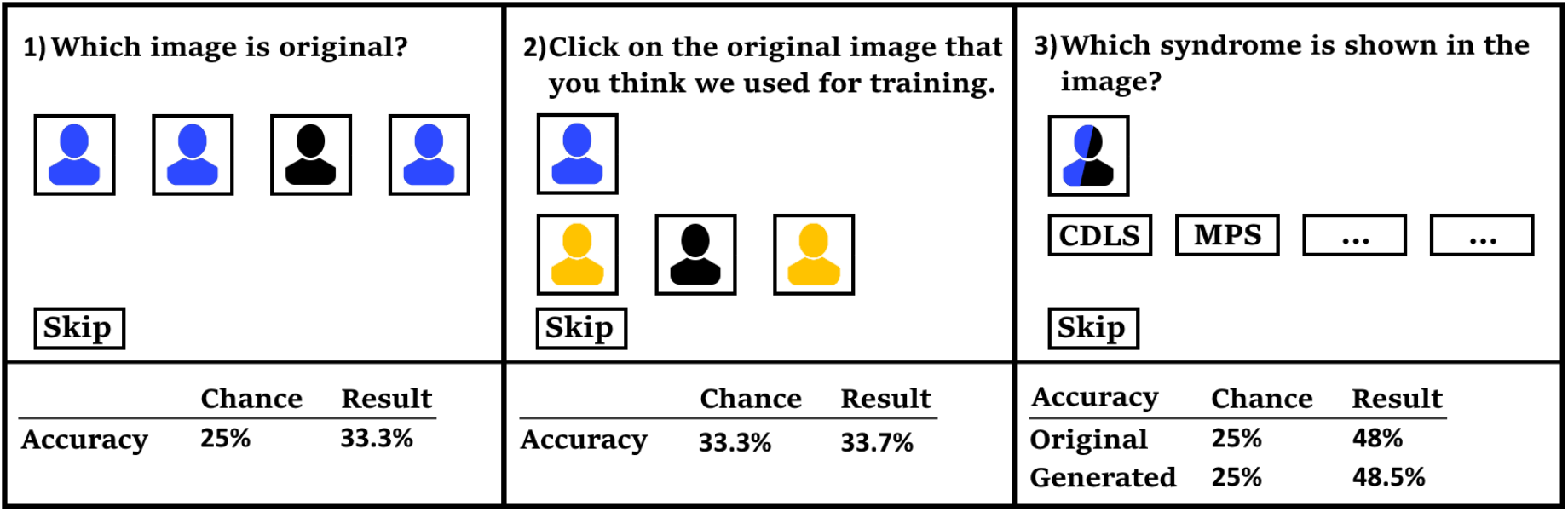
The survey presented to human participants to assess the ability to recognize generated images, specific training images, and specific genetic conditions. The lower section shows the result for each question that was expected due to random chance and what was actually observed. The closer the observed and expected values, the harder is the question. 1) Participants could identify original images slightly more often than randomly expected. 2) Participants could not identify which individuals were used for training. 3) Participants could recognize the characteristic features in original and synthetic images with comparable precision. Color code: original images are depicted in black, original images not part of the training set in yellow, and generated images in blue.

In the first category, participants were presented with four distinct portraits, of which three were generated by GestaltGAN, while one was an image of an individual from the training set. Participants were asked to identify the original. In 33.3% of the cases, participants were able to find the original, exceeding the expected chance value of 25%. The null-hypothesis, that generated images are indistinguishable from real images, had to be discarded (1.9 10-5< 0.05, binomial test). However, this was expected since generated images often contain artifacts that expose them as artificial and still in most cases participants were not able to identify the original.

In the second category, participants were presented with a portrait representing a specific condition, averaged from the latent space of GestaltGAN. The participants were shown three original portraits of individuals with the same condition, only one of which had been used during the training. They were asked to identify the individual who was part of the training set. In 33.7% of cases, participants answered correctly, and the null hypothesis that GestaltGAN generates images that do not violate patient privacy (at least in the tested approach) did hold (0.889> 0.05, binomial test).

In the third category, a synthetic portrait was shown to the participants, and they chose the correct disorder from four different options. If the synthetic images accurately represented the disorders, we hypothesized that experts should be able to identify the correct disorder at approximately the same rate as for real images. Remarkably, in 48% of the cases the experts were able to correctly diagnose the patient based on a real portrait, while their accuracy was 48.5% based on a generated portrait. The null hypothesis, that generated images represent characteristic features comparably to training images, held (0.001 < 0.05, *χ*^2^ test).

## Discussion

In this study, we explored the application of Generative Adversarial Networks in generating photorealistic portraits for rare disorders that preserve the characteristic clinical features but also the patient’s privacy. We presented GestaltGAN, a modified StyleGAN architecture and demonstrated in a series of experiments that synthesizing photorealistic faces of individuals with rare genetic conditions is possible despite limited amounts of training data. Through careful data preparation and augmentation, we were able to generate photorealistic portraits that accurately represent the facial features of a syndrome. Specifically, oversampling and our custom loss function enabled us to train the model to reproduce the characteristic features of disorders more accurately.

Our evaluation encompassed both computational assessments of image quality and human evaluations through an online survey of medical professionals. First, the computational evaluation demonstrated that the majority of generated images were of high-quality, with only a small percentage categorized as fail-cases. The responses of medical professionals further supported this claim and indicated that synthesized portraits represent characteristic features of a given condition and were similar to the original images. Additionally, participants had difficulty in recognizing the original image used for training, suggesting that GestaltGAN can be used to preserve patient privacy.

Using the latent space, we presented the novel latent representations for conditions that average features in the latent space and appear much sharper than simple averages of the faces.

Our study has several important limitations. These include the fact that we assessed a limited number of conditions and focused on a single generative method. While we do not necessarily anticipate that extending the study in these ways would yield very different results, it would be interesting to assess our approach more broadly. In the future, it could be interesting to use more detailed labels, such as additional age labels or individual HPO terms instead of solely the condition in question. This could provide the user more specific control over the generated faces.

In conclusion, we find the parallels of GANs to traditional medical education striking. In medicine, trainees learn medicine according to the mantra of “See one, Do one, Teach one”. Similarly, by training on a relatively small number of cases, GestaltGAN achieved proficiency in generating accurate images of individuals with genetic conditions. The quality of the internalized knowledge was shown as the GAN discriminator, and experts could no longer reliably distinguish artificial and real images. Overall, this work highlights the potential of GANs in the medical field to artificially synthesize data while protecting patient privacy.

## Data Availability Statement

All training data for GestaltGAN was extracted from GMDB. Photorealistic synthetic portraits of 20 disorders can be found at https://thispatientdoesnotexist.org

## Code Availability

We publish our code on github: https://github.com/kirchhoffaron/gestaltgan

## Acknowledgment

This project received institutional funding from Humangenetik Innsbruck.

## Notes

### Competing Interest Statement

The authors have declared no competing interest.

### Funding Statement

This study received institutional funding from University Bonn and University Innsbruck

### Author Declarations

The study used only FAIR data of the GestaltMatcher Database: https://www.medrxiv.org/content/10.1101/2023.06.06.23290887v3 https://db.gestaltmatcher.org/

