## Supplemental_Material for "GestaltGAN: Synthetic photorealistic portraits of individuals with rare genetic disorders"

**Overview of GANs in medical literature**

The creation of artificial images is a complex task, involving teaching the generator to produce realistic images. In GANs, the discriminator evaluates images created by the generator and provides feedback on the image quality. As the generator learns to create more authentic images, the discriminator becomes better at distinguishing between genuine and generated images. In this adversarial process, both improve simultaneously, when the discriminator improves the generator has to synthesize more realistic images to still be able to deceive the discriminator. Since the discriminator provides the loss to the generator, the generator will particularly target features that the discriminator detected as forged during the gradient-descent-based learning. (Goodfellow et al., 2014)

The concept of GANs has been adopted by various architectures such as GigaGAN (Kang et al., 2023) or StyleGAN (Karras, Laine, et al., 2019). StyleGAN has shown promise in artificially generating high-quality images of healthy individuals (i.e., images of individuals in a non-medical context). Throughout the StyleGAN iterations, many of the shortcomings of GANs were addressed, especially artifacts, which can be frequently observed in the generated images.

A notable challenge with GAN’s is mode-collapse, the counterpart to over-fitting in traditional machine learning, where the discriminator inadequately guides the generator, resulting in the generation of a limited set of images, potentially mirroring the original training dataset. Karras, Aittala, Hellsten, et al. (2020) addressed this issue with adaptive discriminator augmentation (ADA)), a method that introduces slight alterations to all images before they are presented to the discriminator. By complicating the classification task for the discriminator, ADA encourages the learning of meaningful rules rather than simply memorizing the original images.

In fields with sparse training data, such as medical imaging, this approach holds particular relevance. GANs have found application in various medical contexts, as demonstrated by Skandarani et al. (2023), who explored their utility for synthesis of liver CT scans and retinal images. Their objective was twofold: to assess the quality of generated images and to investigate their suitability for training other machine learning models, in particular a U-Net for image segmentation. Despite producing high-quality images, they noted that the richness of generated data was inferior to the original training dataset.

Another study by Fang et al. (2023) investigated the generation of Ultra Wide Fundus (UWF) Fluorescein Angiography (FA) photography images. Given the invasive nature of capturing FA images, which requires injecting a potentially harmful contrast agent into the patients’ bloodstream, they sought to transform existing UWF Scanning Laser Ophthalmoscopy images into UWF-FA using GAN. While their model successfully captured many meaningful details such as vascular lesions, it encountered challenges with fine details.

The first study in medical genetics that explored the potential of GANs for next-generation phenotyping (NGP) is from Duong et al. (2022). They used StyleGAN2-ADA to generate images of Williams-Beuren syndrome and 22q11.2 deletion syndrome, focusing particularly on age-related effects on facial features. They successfully controlled the appearance of synthesized faces using age labels, demonstrating the versatility of GANs in medical image synthesis and analysis.

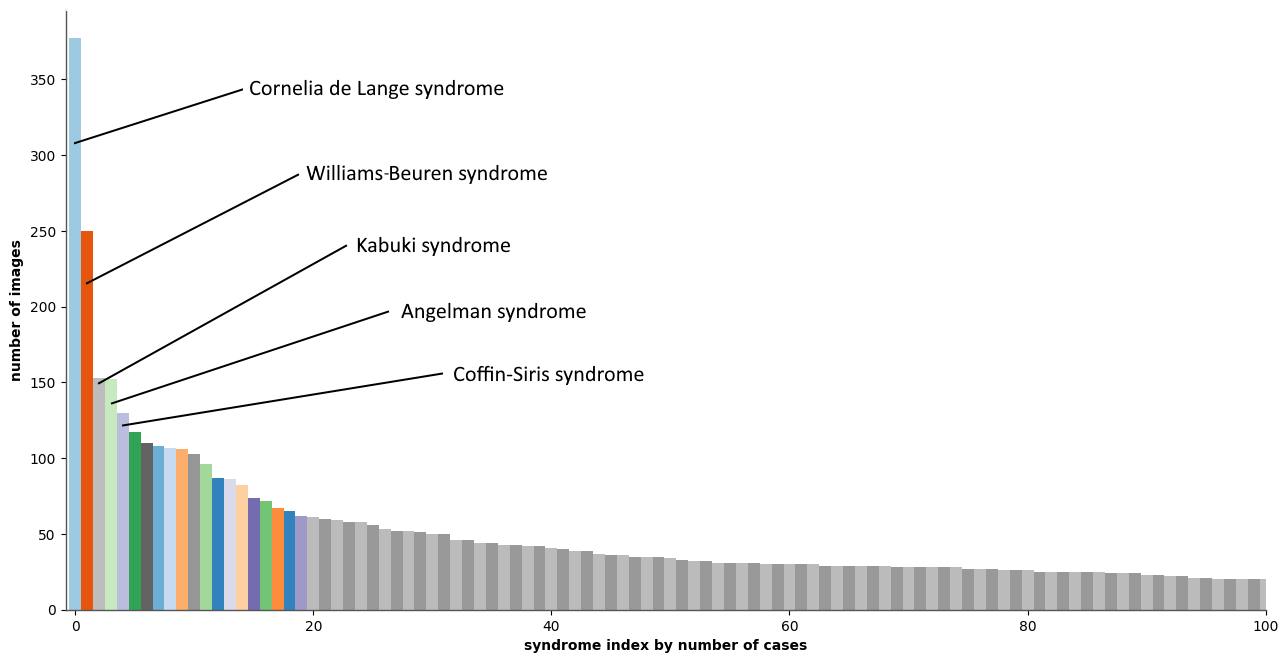

Supplemental Figure 1: Distribution of disorders in the GestaltMatcher database (GMDB). The long tail distribution partially reflects the incidence rates of rare disorders. For instance Cornelia de Lange syndrome Williams Beuren syndrome, Kabuki syndrome, Agnelman syndrome, and Coffin Siris syndrome are more frequent and therefore more cases of these disorders are represented in the GMDB.

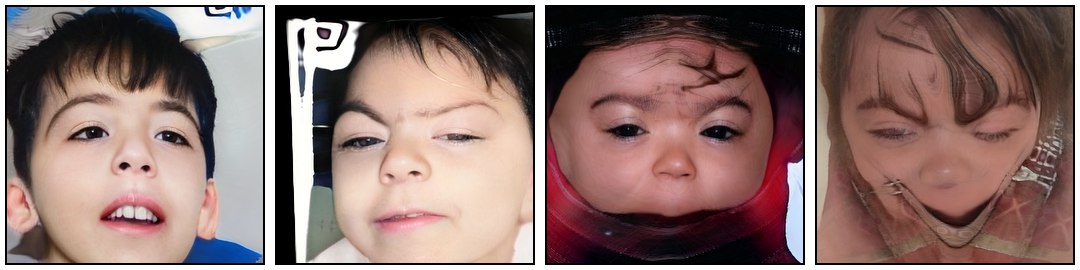

Supplemental Figure 2: Images that achieved a quality score below 99.9% according to RetinaFace and were discarded due to artifacts or missing parts of the face.

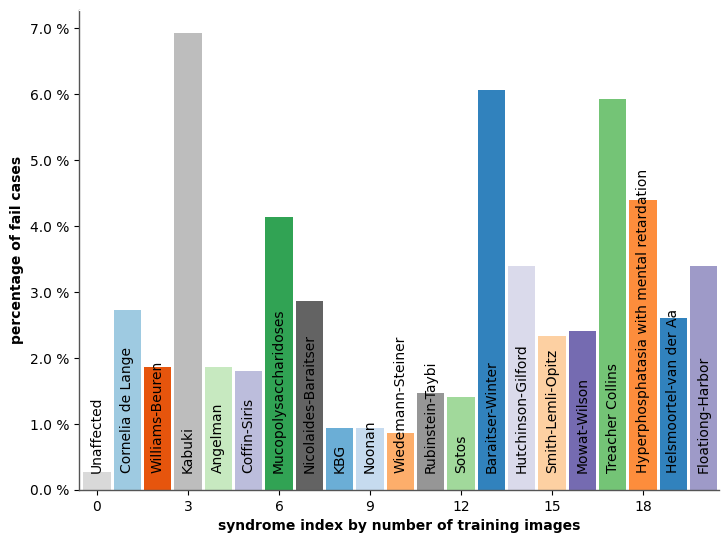

Supplemental Figure 3: Quality control of generated images. RetinaFace was used to assess the quality of a synthetic portrait. A score below 99.9% was chosen as a threshold, since images below this value often missed parts of the eyes, nose, or mouth and would have therefore easily been recognizable as artificial. The proportion of failed cases varied over the disorders.
